# Sharing and Reusing Computable Phenotype Definitions

**DOI:** 10.1101/2023.09.17.23295681

**Authors:** Shyam Visweswaran, Louisa Yu Zhang, Kevin Bui, Eugene M. Sadhu, Malarkodi J. Samayamuthu, Michele M. Morris

## Abstract

**Background:** A scalable approach for the sharing and reuse of human-readable and computer-executable phenotype definitions can facilitate the reuse of electronic health records for cohort identification and research studies.

**Description:** We developed a tool called Sharephe for the Informatics for Integrating Biology and the Bedside (i2b2) platform. Sharephe consists of a plugin for i2b2 and a cloud-based searchable repository of computable phenotypes, has the functionality to import to and export from the repository, and has the ability to link to supporting metadata.

**Discussion:** The i2b2 platform enables researchers to create, evaluate, and implement phenotypes without knowing complex query languages. In an initial evaluation, two sites on the Evolve to Next-Gen ACT (ENACT) network used Sharephe to successfully create, share, and reuse phenotypes.

**Conclusion:** The combination of a cloud-based computable repository and an i2b2 plugin for accessing the repository enables investigators to store and retrieve phenotypes from anywhere and at any time and to collaborate across sites in a research network.

## 1. Introduction

The passage of the Health Information Technology for Economic and Clinical Health (HITECH) Act by the United States federal government accelerated the adoption of electronic health records (EHRs) (1, 2). This widespread adoption of EHRs spurred the reuse of EHR data to advance biomedical research and discovery. Broadly speaking, EHRs contain structured or coded data, free text data, and imaging data, of which structured data that consists of patient information documented using controlled vocabularies is widely used in research. A key component of the reuse of EHR data is the development, sharing, and application of computable phenotype definitions (computable phenotypes for short) that are computer-executable definitions to identify patients with specific criteria (3, 4). Computable phenotypes enable reproducible queries of EHR data in a high throughput fashion both within and across healthcare organizations and are particularly useful in the identification of cohorts of people with specific conditions. Phenotype definitions are composed of data elements and logic expressions (AND, OR, NOT) that can be interpreted and executed by a computer, and standardization of the definitions rely on terms and value sets of controlled vocabularies used in coding EHR data. Due to variabilities in healthcare processes, coding and reimbursement practices, experience of providers, access to healthcare, data in EHRs are often incomplete and biased. Thus, developing and validating computable phenotypes of high accuracy is often time-consuming and resource intensive. Further, the lack of approaches and tools for sharing and reuse inhibits the widespread use of computable phenotypes within and across healthcare organizations.

In this paper, we describe the development of a tool, Sharephe, for the sharing and reuse of computable phenotype definitions for the Informatics for Integrating Biology and the Bedside (i2b2) platform (5). Sharephe (pronounced Sheriff) provides a cloud-based searchable repository of computable phenotypes, tools to import to and export from the cloud-based repository, and the ability to link to supporting metadata. This tool can enable clinical research networks to realize large-scale sharing and reuse of human-readable and computer-executable phenotype definitions, thereby advancing data-driven precision medicine research. While Sharephe is currently limited in its application to the i22b platform, it provides a foundation to expand to other platforms.

## 2. Background

Patient cohort identification is the identification of patients who satisfy predefined criteria from a large population and is a necessary step in clinical trial recruitment, translational and retrospective research studies, quality improvement studies, and the creation of disease registries. Much of cohort discovery is a time-consuming and expensive process that is done by research staff by querying disparate clinical systems and reviewing medical charts for patients matching a specific set of criteria.

The widespread deployment of EHRs has provided the opportunity for computationally exploring and processing patient data. The use of EHR data for cohort identification and, more generally, the reuse of EHR data necessitates the transformation of the data into an easily analyzed format. In response to this challenge, many healthcare organizations have established clinical data warehouses, which are specialized databases that integrate data from disparate clinical systems to facilitate the reuse of data. Often, these warehouses use custom-designed architectures and formats, and accessing the data in them needs specialized knowledge of the database architecture and database query languages such as SQL. To overcome these obstacles, a number of solutions have emerged, such as the introduction of common data models to facilitate collaboration across healthcare organizations and the emergence of user interfaces to facilitate the creation of queries without the need to know database query languages.

The advent of common data models has enabled federating research analyses and aggregation of the results. The OMOP Common Data Model is a widely used common data model that is provided by the Observational Medical Outcomes Partnership (OMOP) and is used to harmonize longitudinal patient data with standardized medical terminologies (6). PCORnet, the National Patient-Centered Outcomes Research Network, developed the PCORnet Common Data Model, which is used to integrate data to enable large-scale comparative effectiveness research (7). Another advance is the use of interfaces with structured terms that are used to construct complex queries. For example, i2b2 is a clinical data warehouse platform that has been widely implemented and employs an ontology-based query interface that allows easy creation of complex computable phenotypes (5). i2b2 is widely utilized both as a standalone warehouse and in federated networks, such as the Accrual of Patients to Clinical Trials (ACT) network and its successor, Enable Next-Gen ACT (ENACT) network (8).

Typically, computable phenotypes are described in human-readable documents as a set of inclusion and exclusion criteria with logical constraints (e.g., AND, OR, NOT) with additional temporal constraints (9). These descriptions are then translated into a specific query language for execution in the clinical data warehouse. The advent of research networks has enabled the sharing and reuse of computable phenotypes to an extent. For example, the Electronic Medical Records & Genomics (eMERGE) network has developed more than 50 phenotypes that use billing codes, laboratory test results, medication data, and natural language processing of clinical notes (10), and the Pharmacogenomics Research Network (PGRN) has developed drug-response phenotypes (11) that have been shared across multiple sites. Furthermore, the widespread use of common data models such as OMOP, PCORnet, and ENACT has enabled the authoring of an executable phenotype that is reusable with almost no modifications at multiple institutions. However, the sharing of queries is an internal process in these networks that is not necessarily standardized or scalable. Though a knowledge repository of phenotypes exists – the Phenotype Knowledge Base (PheKB) (12) – that hosts descriptions and definitions of phenotypes from a range of projects such as eMERGE, PCORnet, and PGRN, readily executable phenotypes are not widely available for reuse.

A scalable approach for sharing and reuse of human-readable and computer-executable phenotypes can facilitate extensive reuse of EHR data for cohort identification and other uses. Moreover, widespread sharing and reuse of phenotypes will improve standardization and validation of the phenotype definitions. Richesson et al. identified four components for informed sharing and reuse that include 1) searchable libraries of computable phenotypes, 2) knowledge bases with information and methods; 3) tools to create, execute, share, and reuse phenotypes; and 4) motivated users and stakeholders (13).

## 3. Description of Sharephe

The Sharephe tool consists of an i2b2 plugin, a cloud-based repository, and a website. We first describe the user functionality of Sharephe and then briefly describe the technical architecture of the tool.

### 3.1 User Functionality

Once installed, the Sharephe plugin is accessible to the user from the i2b2 interface, and it provides the following functions.

#### A. Access searchable repository

As shown in Figure 1, the “Library” tab in Sharephe enables the user to connect to the cloud-based repository, search for specific phenotypes, and browse available phenotypes. In addition to the plugin, the Sharephe website enables any user to browse and search the repository for phenotypes (see Figure 2).

**Figure 1.**
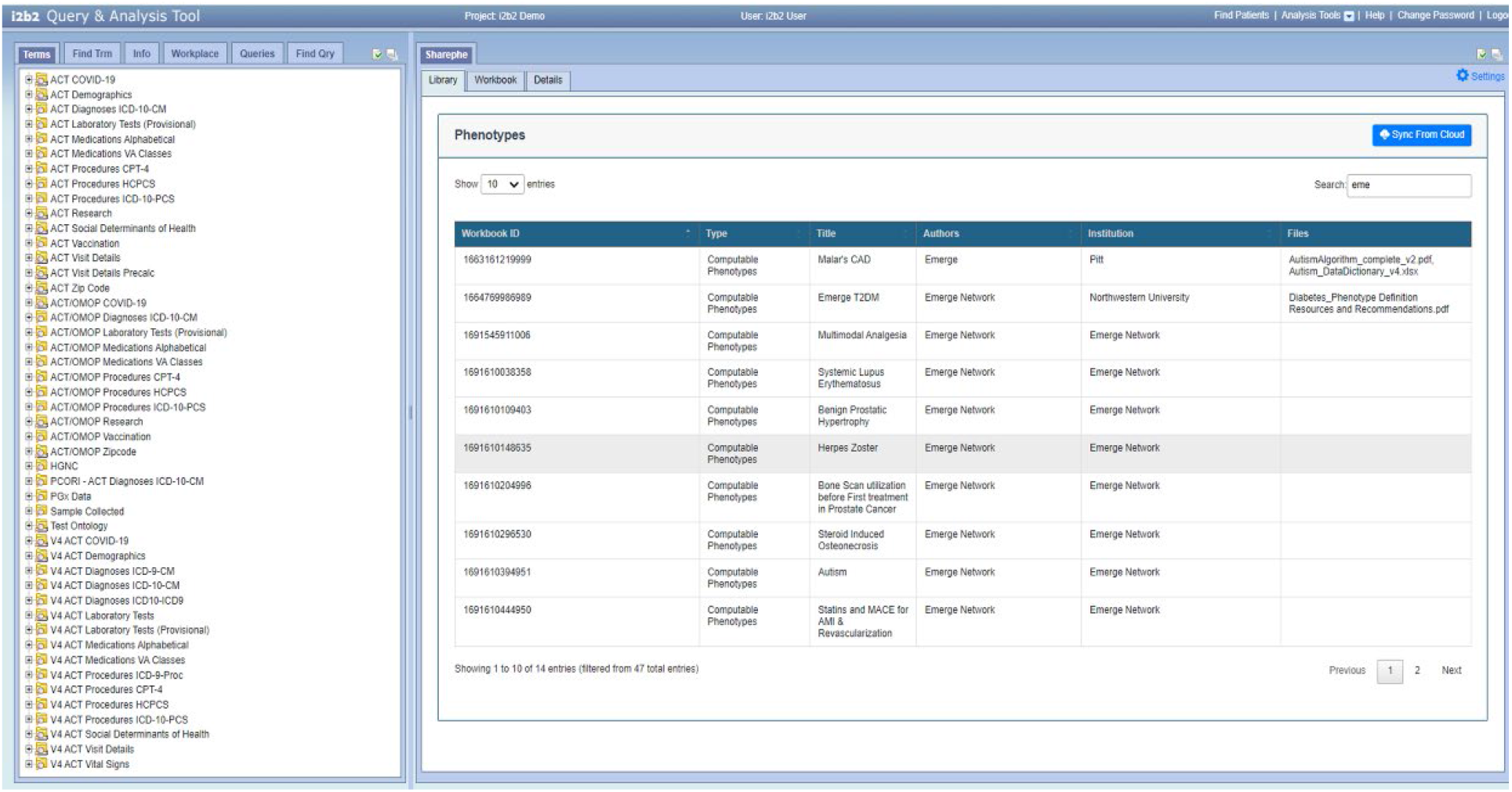
Sharephe plugin displaying phenotypes in the cloud-based repository.

**Figure 2.**
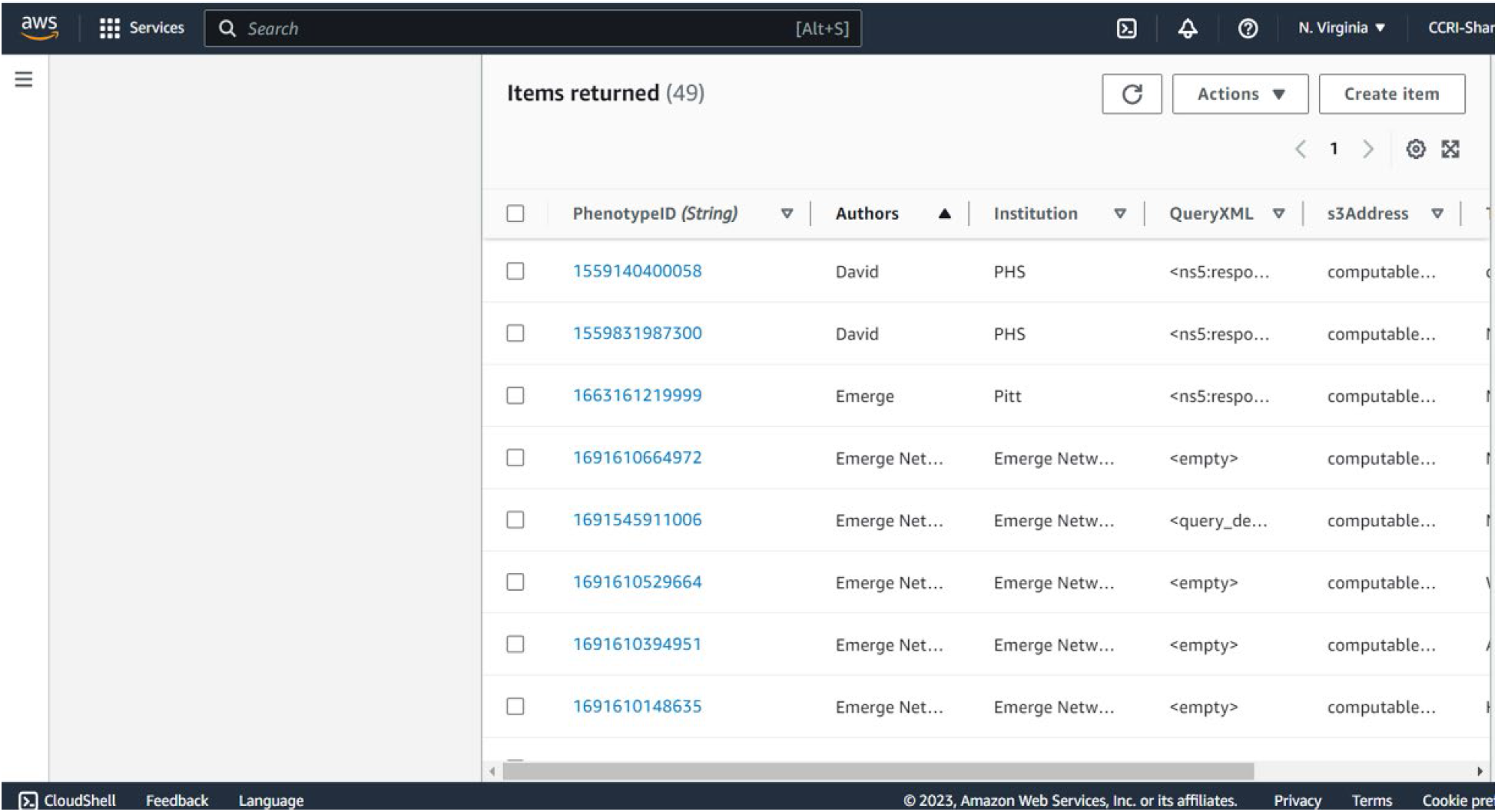
Sharephe website displaying phenotypes in the cloud-based repository.

#### B. Import phenotype workbook

In the cloud-based repository, phenotypes are organized into workbooks, each of which contains a collection of executable phenotypes. Figure 3 illustrates a workbook on autism that contains five phenotypes, with the first three defining cases (Autistic Disorder, Asperger’s Syndrome, and PDD NOS) and the next two defining controls (Control 1, Control 2). A user can select and import (download) one or more relevant workbooks. The description of the workbook and its constituent queries are displayed in the “Description” tab after importing a workbook (see Figure 3). Currently, a Sharephe workbook consists of a unique identifier, up to ten phenotypes, user and institution metadata, and an optional set of associated objects.

**Figure 3.**
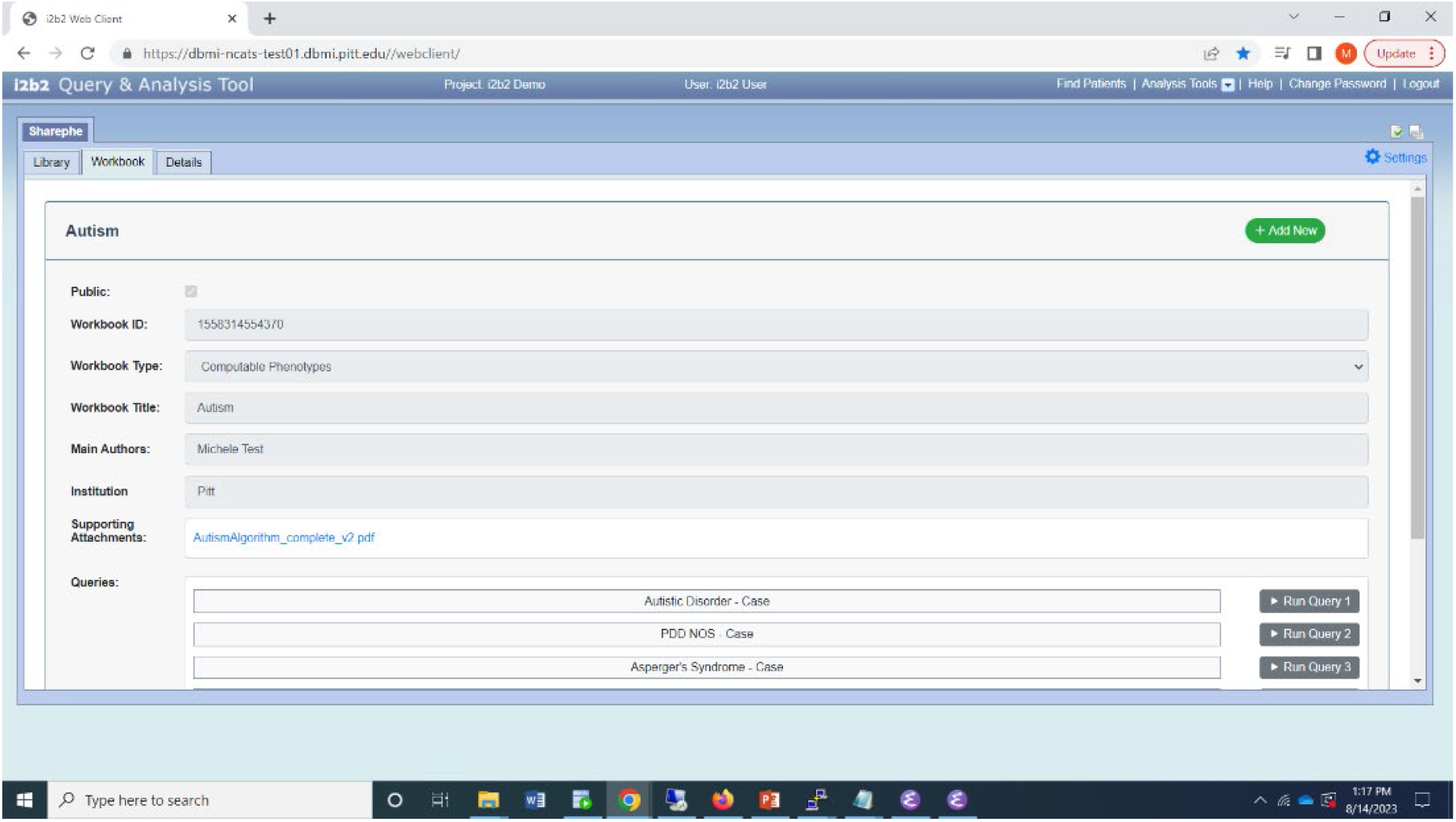
Sharephe plugin displaying a workbook.

#### C. Execute phenotypes

After importing a workbook, the user can review associated information, including descriptions, author-provided instructions, and related publications. Each phenotype in the workbook has its own execute button, allowing the user to execute the phenotype on the local i2b2 data mart (see Figure 3). Sharephe uses the standard i2b2 CRC cell to execute each phenotype, and once the phenotype has been executed, the “Find Patients” panel displays patient counts, and the phenotype is saved in the “Previous Queries” panel. The user can subsequently execute or modify these phenotypes.

#### D. Export phenotype workbook

The ‘Description’ tab also permits the creation of phenotypes for uploading to the cloud-based repository. To create a new workbook, the user must click the “New” button and enter the necessary information. To define phenotypes, the user drags and drops one or more existing queries from the local i2b2 onto the panel. This workbook can then be uploaded as a Sharephe workbook to the repository.

#### E. Obtain phenotype details

The “Details” tab displays a human-readable list of the terms in the query as well as the logical and other operations that are defined in the query (see Figure 4).

**Figure 4.**
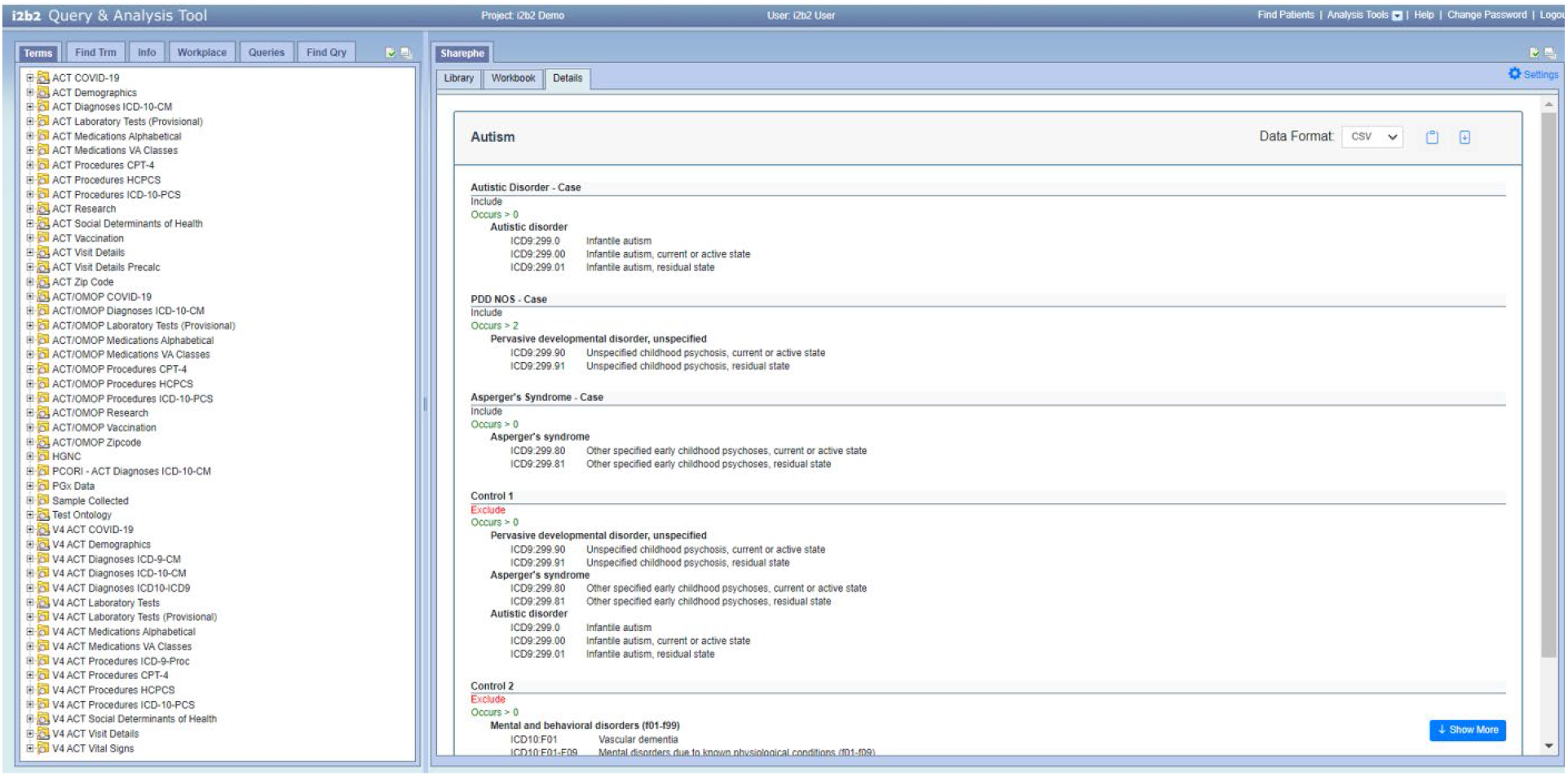
Sharephe plugin displaying details of a query.

### 3.2 Technical Architecture

The i2b2 platform has a modular “hive” architecture with a set of server-side modules called “cells” and a web client system that is implemented in HTML and JavaScript (14). The Sharephe plugin follows the i2b2 architecture and implements two server-side “cells” that are called SharepheCloud and SharepheOntology (see Figure 5).

**Figure 5.**
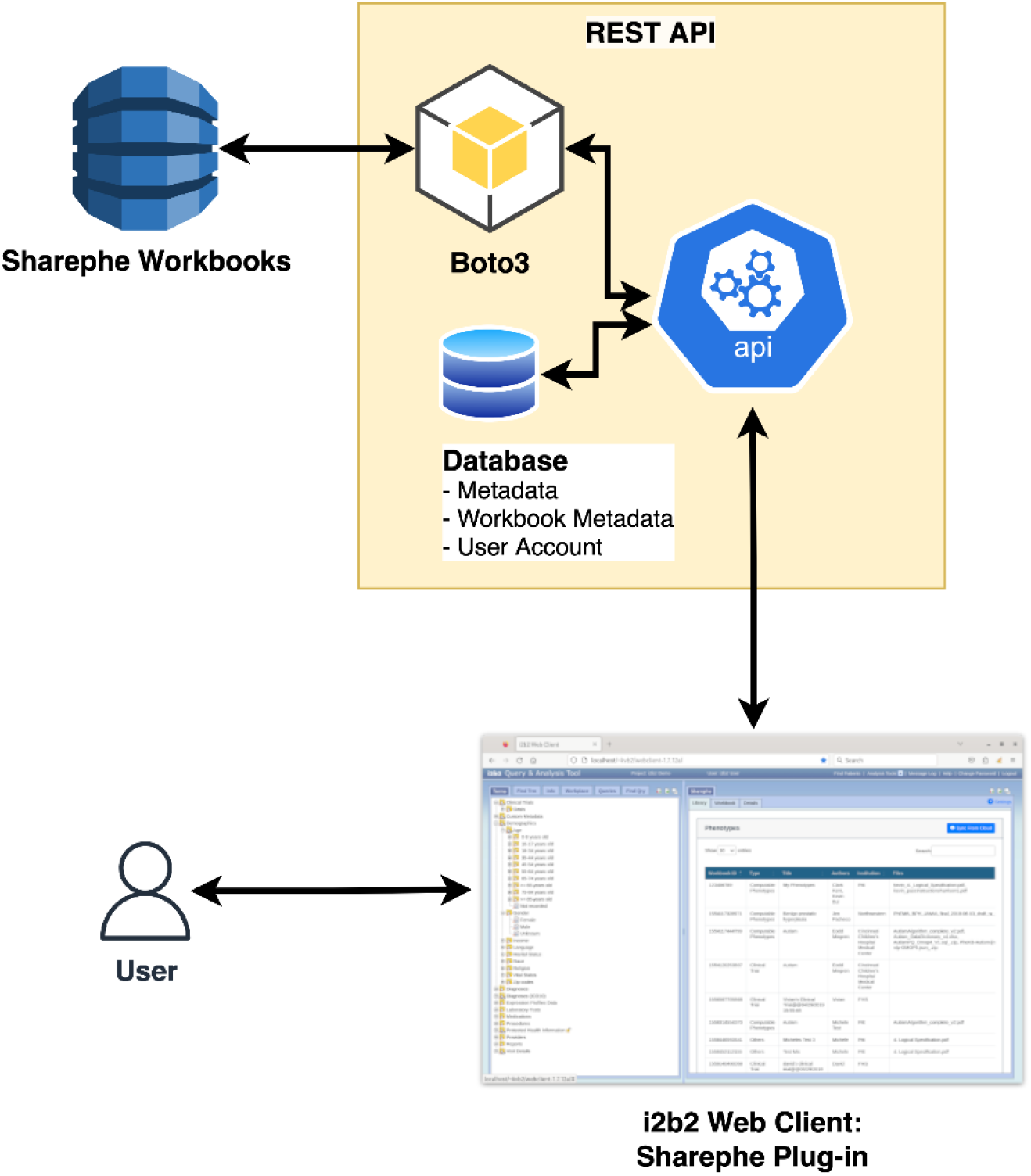
Architecture of the Sharephe plugin.

The primary cell SharepheCloud is the connection between the i2b2 server and the Amazon AWS cloud repository. SharepheCloud employs Amazon’s BasicAWSCredentials interface, which enables callers to authenticate using the AWS access key and secret access key. It then employs AmazonDynamoDBClient and AmazonS3ClientBuilder to transfer data between a user’s local i2b2 and the repository on AWS’s cloud. DynamoDB is a Non-SQL database in which the Sharephe workbook’s structured elements are stored. Currently, the workbook contains a collection of i2b2-formatted XML queries, a list of authors, institutions, the title of the phenotype, and the query in human-readable format. Sharephe employs Amazon Simple Storage Service through a secure web service to store objects such as images, Word documents, and PDFs that support the computable phenotype in the repository. These objects are linked to the corresponding DynamoDB structured data. S3 also supports data management at the account, bucket, and object levels, enabling Sharephe to manage user roles and access.

Sharephe also extends the capabilities of the i2b2 core Ontology Cell in a new Sharephe Ontology cell. This cell contains methods for traversing the ontology, translating the query into a human-readable format, and listing the phenotype’s terms and their sources.

### 3.3 Website

In addition to the i2b2 plugin, Sharephe includes a complementary website. Non-i2b2 users are able to browse the phenotype library and download executable versions of the phenotypes via the website. The Sharephe plugin and the Sharephe library are available at http://www.sharephe.org.

### 3.4 Preliminary Assessment

We locally evaluated Sharephe by implementing the autism workbook, which contained three case phenotypes and two control phenotypes obtained from PheKB. Included in the autism workbook’s metadata were the original authors and supporting documentation. The autism workbook was exported from one i2b2 installation to the cloud-based repository and imported into a second i2b2 installation, where it was successfully executed to obtain patient counts. Additionally, we tested the tool’s functionality at two ENACT sites. The Sharephe tool was successfully installed at a second site, and the autism workbook was successfully shared and reused between the two sites.

## 4. Discussion

National initiatives like the All of Us Research Program (15), and research networks like ENACT, PCORnet, eMERGE, and PGRN rely on high throughput computable phenotyping that are shared widely and regularly reused. Validating, standardizing, and iteratively refining computable phenotypes in these initiatives and networks requires widespread and effortless sharing.

As a step toward enabling the sharing and reuse of computable phenotypes on the i2b2 platform, we developed the Sharephe tool. In the context of the ENACT network, which has deployed i2b2 data marts with a common ontology at over 55 sites, Sharephe can leverage the common technology, data model, and ontology to facilitate the sharing of phenotypes. Sharephe’s cloud-based repository provides searchable libraries of phenotypes as well as information describing them and executable queries that can be downloaded and executed on an i2b2 system with a compatible ontology. The i2b2 platform enables researchers to create, evaluate, and implement phenotypes without knowing complex query languages. The ENACT network’s shared ontology ensures that queries developed at one site will be executed without modification at any other site. In addition, Sharephe’s functionality permits the export and import of phenotypes individually or in bulk. A key objective of the ENACT network is to identify patients at multiple sites to facilitate multi-site clinical trials and for research studies; this objective aligns with the users’ desire to share and reuse phenotypes within the network.

### 4.1 Limitations and Future Work

Even though Sharephe can facilitate the sharing of phenotypes on a large national research network, it has several limitations. Currently, phenotypes are only executable on the i2b2 platform; however, we intend to provide automatic translations of i2b2 queries into executable implementations for the OMOP and PCORnet common data models in future work. Current phenotypes are incompatible if sites use different versions of the ENACT ontology. We are investigating methods for automatically adapting phenotypes constructed in one version of an ontology to execute in a different version.

## 5. Conclusions

Sharephe is a tool for sharing and reusing human-readable and computer-executable phenotypes on the i2b2 platform. In addition, it permits rapid archiving and sharing of phenotypes via a cloud-based repository. The combination of a cloud-based repository and an i2b2 plugin for accessing the repository enables investigators to store and retrieve phenotypes from anywhere and at any time, and to collaborate across sites.

## Data Availability

All data produced are available online at http://www.sharephe.org.

## Acknowledgments

The research reported in this publication was supported by the National Center for Advancing Translational Sciences of the National Institutes of Health under award numbers UL1 TR001857 and U24 TR004111. The content is solely the responsibility of the authors and does not necessarily represent the official views of the National Institutes of Health. The authors thank Desheng Li for technical assistance.

## Notes

### Competing Interest Statement

The authors have declared no competing interest.

## References

1. Blumenthal D. Launching HITECH. New England Journal of Medicine. 2010;362(5):382–5.

2. Blumenthal D. Stimulating the adoption of health information technology. New England Journal of Medicine. 2009;360(15):1477–9.

3. Pathak J, Kho AN, Denny JC. Electronic health records-driven phenotyping: challenges, recent advances, and perspectives. BMJ Publishing Group BMA House, Tavistock Square, London, WC1H 9JR; 2013.

4. Richesson RL, Sun J, Pathak J, Kho AN, Denny JC. Clinical phenotyping in selected national networks: demonstrating the need for high-throughput, portable, and computational methods. Artificial Intelligence in Medicine. 2016;71:57–61.

5. Murphy SN, Weber G, Mendis M, Gainer V, Chueh HC, Churchill S, et al. Serving the enterprise and beyond with informatics for integrating biology and the bedside (i2b2). Journal of the American Medical Informatics Association. 2010;17(2):124–30.

6. Hripcsak G, Duke JD, Shah NH, Reich CG, Huser V, Schuemie MJ, et al. Observational Health Data Sciences and Informatics (OHDSI): opportunities for observational researchers. Studies in Health Technology and Informatics. 2015;216:574.

7. Fleurence RL, Curtis LH, Califf RM, Platt R, Selby JV, Brown JS. Launching PCORnet, a national patient-centered clinical research network. Journal of the American Medical Informatics Association. 2014;21(4):578–82.

8. Visweswaran S, Becich MJ, D’Itri VS, Sendro ER, MacFadden D, Anderson NR, et al. Accrual to Clinical Trials (ACT): A Clinical and Translational Science Award Consortium Network. JAMIA open. 2018;1(2):147–52.

9. Richesson RL, Hammond WE, Nahm M, Wixted D, Simon GE, Robinson JG, et al. Electronic health records based phenotyping in next-generation clinical trials: a perspective from the NIH Health Care Systems Collaboratory. Journal of the American Medical Informatics Association. 2013;20(e2):e226–e31.

10. Denny JC, Bastarache L, Ritchie MD, Carroll RJ, Zink R, Mosley JD, et al. Systematic comparison of phenome-wide association study of electronic medical record data and genome-wide association study data. Nature Biotechnology. 2013;31(12):1102.

11. Shuldiner A, Relling M, Peterson J, Hicks K, Freimuth R, Sadee W, et al. The pharmacogenomics research network translational pharmacogenetics program: overcoming challenges of real-world implementation. Clinical Pharmacology & Therapeutics. 2013;94(2):207–10.

12. Kirby JC, Speltz P, Rasmussen LV, Basford M, Gottesman O, Peissig PL, et al. PheKB: a catalog and workflow for creating electronic phenotype algorithms for transportability. Journal of the American Medical Informatics Association. 2016;23(6):1046–52.

13. Richesson RL, Smerek MM, Cameron CB. A framework to support the sharing and reuse of computable phenotype definitions across health care delivery and clinical research applications. eGEMs. 2016;4(3).

14. Mendis M, Wattanasin N, Kuttan R, Pan W, Philips L, Hackett K, et al., editors. Integration of Hive and cell software in the i2b2 architecture. In: Proceedings of the AMIA Annual Sympoisum; 2007.

15. Collins FS, Varmus H. A new initiative on precision medicine. New England Journal of Medicine. 2015;372(9):793–5.

